# African ancestry *APOE* e4 non-carriers with higher educational attainment are resilient to Alzheimer disease pathology-specific blood biomarker pTau181

**DOI:** 10.1101/2023.07.06.23292263

**Authors:** Farid Rajabli, Azizi A. Seixas, Bilcag Akgun, Larry D. Adams, Jovita Inciute, Takiyah Starks, Renee Laux, Goldie S Byrd, Jonathan L Haines, Gary W Beecham, Jeffery M Vance, Michael L Cuccaro, Margaret A. Pericak-Vance

## Abstract

Cognitive and functional abilities in individuals with Alzheimer disease (AD) pathology (ADP) show greater than expected variability. While most individuals show substantial impairments in these abilities, a considerable number show little or no impairments. Factors contributing to this variability are not well understood. For instance, multiple studies have shown that higher levels of education are associated with reduced cognitive impairments among those with ADP. However, it remains unclear whether higher levels of education are associated with functional impairments among those with ADP.

We studied 410 AA individuals with advanced levels of pTau181 (a biomarker for ADP; individuals as those having log_10_(pTau181) level greater than one standard deviation above the mean) to determine whether EA (categorized as low EA for individuals with ≤ 8 years of education and high EA for those with >8 years) promotes functional resilience and whether this effect varies between APOE ε4 carriers and non-carriers. We used the four non-memory components of the Clinical Dementia Rating (CDR) to create a composite score (CDR-FUNC) to evaluate functional difficulties (scored from 0=no impairment to 12=severe). We employed the non-parametric Mann-Whitney U test to assess the relationship between EA and CDR-FUNC in advanced levels of pTau181 individuals.

The results showed that EA promotes resilience to functional problems in AA individuals with advanced levels of pTau181, such that individuals with high EA are more likely to have better functional ability compared to those with lower EA (W=730.5, p=0.0007). Additionally, we found that the effect of high EA on functional resilience was stronger in ε4 non-carriers compared to ε4 carriers (W=555.5, p=0.022).

This study extends the role of cognitive reserve and EA to functional performance showing that cognitive reserve influences the association between ADP burden and functional difficulties. Interestingly, this protective effect seems less pronounced in carriers of the strong genetic risk allele ε4. The results highlight the intricate interplay of genetic and non-genetic factors in AD progression, suggesting a need for more personalized strategies to manage functional decline in AD.

## BACKGROUND

Alzheimer disease (AD) is the most common type of dementia, with older African ancestry individuals in US having almost 2 times higher prevalence and incidence compared with older non-Hispanic Whites (NHW) (Folstein et al., 1991; Gurland et al., 1999, Alzheimer Association Report). Several factors influence the incidence, prevalence, and risk of AD. These include genetic, clinical, social, behavioral, and environmental determinants (Gatz et al., 2006; Migliore and Coppedè, 2022; Litke et al., 2021; Majoka and Schimming, 2021; Nianogo et al., 2022; Bellenguez et al. 2022).

The characteristic neuropathological changes of AD, the presence of extracellular amyloid-beta deposits and intracellular aggregation of neurofibrillary tangles, can now be measured using biomarkers (Jack et al., 2018; Hampel et al., 2021). These include neuroimaging, cerebrospinal fluid (CSF), and blood-based biomarkers (Jack et al., 2018; Hampel et al., 2021). Recent developments in the field of blood biomarkers showed that blood pTau181 levels strongly correlate with abnormal amyloid accumulation, tau deposition, and neurodegeneration (Karikari et al., 2020).

ADP is typically associated with the decline in cognitive, functional, and behavioral abilities seen in AD (Hampel et al. 2021). However, some studies have shown that even with advanced ADP, a subgroup of individuals can still maintain reasonable cognition and function (Arenaza-Urquijo and Vemuri, 2018; Stern et al., 1994). This inconsistency between ADP and clinical symptoms underscores a possible phenotype characterized by biological pathology without clinical impairment.

Research indicates that a concept called ‘cognitive reserve’ (CR) could provide this protective barrier against cognitive impairment in the presence of ADP (Stern and Barulli, 2019; Stern, 2012). CR is thought to be the various thinking abilities that actively compensate for the deficits imposed by the ADP (Stern et al., 2019). This compensation yields cognitive performance that exceeds expectations when considering life-course-related brain changes and brain injuries or disease (Collaboratory on Research Definitions for Reserve and Resilience in Cognitive Aging and Dementia, 2022).

Educational attainment (EA), a commonly used indicator for CR, significantly influences cognitive performance among individuals with similar levels of ADP. Higher EA tends to foster resilience against AD clinical manifestations, often resulting in improved cognitive performance compared to those with less education (Avila et al., 2021; Lovden et al., 2020; Seblova et al., 2020; Stern et al., 2020). Interestingly, studies suggest an intricate interplay between EA and the ***APOE*** ε4 allele, the strongest genetic risk factor for late-onset AD (Corder et al., 1993; Vonk et al., 2019; Dekhtyar et al., 2019). This complex relationship indicates that CR and genetic factors might interact and influence cognitive outcomes. However, it remains unclear how this protective effect of CR translates into functional abilities, such as managing finances, social interactions, and personal care and whether this protection influenced by ***APOE*** gene.

In this study, we investigated whether EA, a proxy for cognitive reserve, promotes functional resilience in African ancestry individuals with advanced levels of pTau181 (used as a proxy for ADP) and whether this effect differs between carriers and non-carriers of the ***APOE***-ε4 allele.

## METHODS

### Study Samples

We ascertained 410 participants through AD genetics studies from Wake Forest University (WF, North Carolina), and University of Miami (UM, Florida). All participants were classified Black American based on self-reported race-ethnicity. The inclusion criteria for the study were having CDR scores, reported years of education, *APOE* genotypes, and blood pTau181 AD biomarker data. All participants or their consenting proxy provided written informed consent as part of the study protocols approved by the site-specific Institutional Review Boards.

Individual ancestral backgrounds were confirmed using genome-wide genetic data with EIGENSTRAT software (Price et al., 2006). Population substructure data sets were compared with those in the 1000 Genome reference panel YRI (Yoruba from Nigeria) and CEU (Utah Residents with Northern and Western European ancestry) populations. Outliers with respect to CEU population (overlapping within the cluster of CEU) were removed from the datasets (1000 Genomes Project Consortium et al., 2015).

Ascertainment protocols have been consistent across the sites and clinical data assessments capture sociodemographic information including years of education, medical and family history, dementia staging, AD/dementia symptoms, neuropsychological abilities, functional capabilities, and behavioral impairments. The CDR was used to evaluate functional capabilities and served as an outcome measure as described below in the Statistical Analysis section. Biomaterials were also collected at the time of study entry by trained phlebotomists. The detailed description of the study samples, recruitment and cognitive assessment are described elsewhere (Vonk et al., 2019).

#### Genotyping

Genome-wide single-nucleotide polymorphism (SNP) genotyping was processed on Global Screening Array and *APOE* genotyping was performed as described in Saunders et al (Saunders et al. 1993).

### Measurement of Serum AD Biomarkers pTau181

Serum concentrations of pTau181 were measured using SIMOA chemistry implemented on the Quanterix HD-X instrument (Quanterix, Billerica, MA, USA){{52699 Wilson,D.H. 2016}} according to manufacturer’s instructions for the pTau181 Advantage V2 assay (catalog #103714). Samples were randomized according to age, sex, and diagnosis and assayed in duplicate on each plate. Biomarker levels were log10-transformed to satisfy normality assumptions. Samples were removed if biomarker levels were greater than three standard deviations from the mean of the rest of the samples.

### CDR-FUNC

The Clinical Dementia Rating scale (CDR) is a semi-structured interview that assesses cognitive and functional impairment associated with AD to allow for staging based on a global CDR score (Hughes et al. 1982). The global CDR score is derived from box scores assigned to six domains: Memory, Orientation, Judgment and Problem solving, Home and Hobbies, Community Involvement, and Personal Care. Various studies have examined the utility of these box scores for clinical and research purposes (Julayanont et al., 2022, O’Bryant et al., 2008; Grober et al., 2021). The application of box scores in the assessment of functional impairment has also been studied. For example, Sudo et al. developed a composite score designed to assess functional difficulties (CDR-FUNC) (Sudo et al., 2016). This score is based on the sum of CDR box scores that are aligned with functional performance (Problem solving, Home and Hobbies, Community Involvement, and Personal Care) and which may have a different trajectory than CDR based cognitive abilities. In addition, Sudo and colleagues reported that the CDR-FUNC score is strongly correlated with the functional performance as measured by the Pfeffer’s Functional Activities Questionnaire (FAQ) (Sudo et al., 2016; Pfeffer et al., 1982).

### Statistical Analysis

We hypothesized that individuals with high EA and advanced levels of pTau181 would present better-than-expected functional difficulties as compared to their counterparts with low EA. We further investigated whether EA promotes functional resilience (abilities managing money, interacting with others, and caring for personal needs) differently between *APOE ε4* allele carriers and non-carriers. To model our hypothesis, we used EA as a proxy for cognitive reserve, CDR-FUNC as a measure for functional difficulties, and pTau181 as a proxy for ADP. We categorized and quantified the variables as described below:

#### EA

We stratified years of education into two categories: low EA, individuals up to high school (≤ 8 years), and high EA, individuals beyond high school (>8 years) (Vonk et al. 2019).

#### CDR-FUNC

Using the non-memory components of the CDR, Judgment and Problem solving, Home and Hobbies, Community Involvement, and Personal Care, we formulated a composite score of 12, totaling the individual score of the 4 components with the range of 0 to 3 (Sudo et al. 2016). The CDR-FUNC score ranges from 0 to 12, with lower numbers indicating no to mild functional difficulties and higher scores indicating more severe difficulties.

#### ADP (pTau181)

We defined advanced ADP individuals as those having log_10_(pTau181) level greater than one standard deviation above the mean.

#### Statistical test

To examine the relationship between EA and CDR-FUNC in individuals with advanced ADP, we used the non-parametric Mann Whitney U Test (MW). First, we conducted the analysis using the entire sample of advanced ADP individuals. Next, to determine whether the protective effect of high EA on functional resilience is modified by the presence of the ε4 allele, a genetic risk factor for AD, we stratified our sample by ε4 status (carriers and non-carriers) and re-analyzed the association between EA and CDR-FUNC in each subgroup.

## RESULTS

The study sample consisted of 73.9% females, with a mean age of 71.08 (SD=7.42) years and a mean log10(pTau181) level of -0.01 (SD=0.42) (Table 1). A total of 74 individuals were classified as having advanced log_10_(pTau181) level (mean+1SD), with 43 carrying the ***APOE*** ε4 allele and 31 not carrying it. Individuals with advanced log_10_(pTau181) and low EA had significantly higher CDR-FUNC scores (median: 12.0; Q1: 8.0 - Q3: 12.0) compared to those with high EA (median: 3.75; Q1: 0 - Q3: 9.0; W=730.5, p=0.0007). The distribution of CDR-FUNC scores between low and high EA in individuals with advanced log_10_(pTau181) is illustrated in Figure 1A.

**Table 1.**
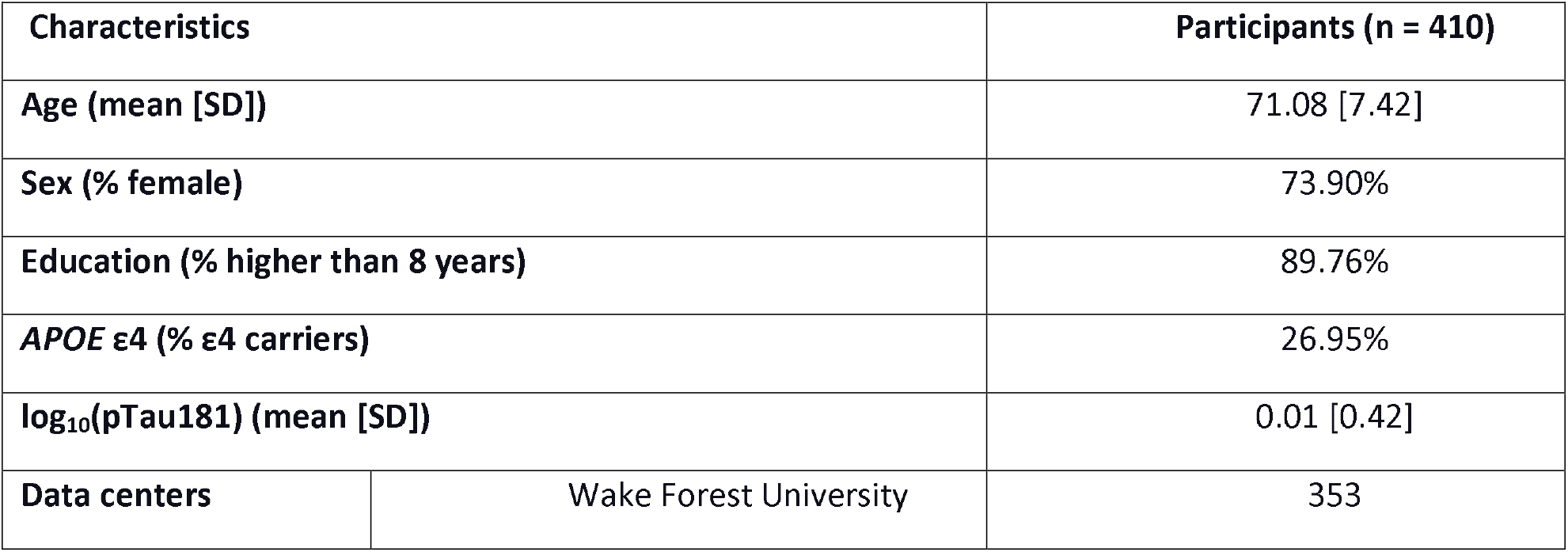
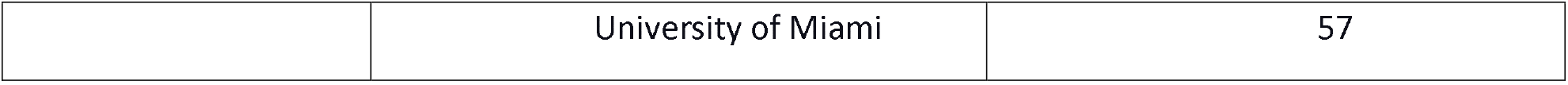
Study samples characteristics

| Characteristics |  | Participants (n = 410) |
| --- | --- | --- |
| Age (mean [SD]) |  | 71.08 [7.42] |
| Sex (% female) |  | 73.90% |
| Education (% higher than 8 years) |  | 89.76% |
| *APOE* $\epsilon 4$ (% $\epsilon 4$ carriers) | | 26.95% |
| $\log_{10}(\text{pTau181})$ (mean [SD]) | | 0.01 [0.42] |
| Data centers | Wake Forest University | 353 |

**Figure 1.**
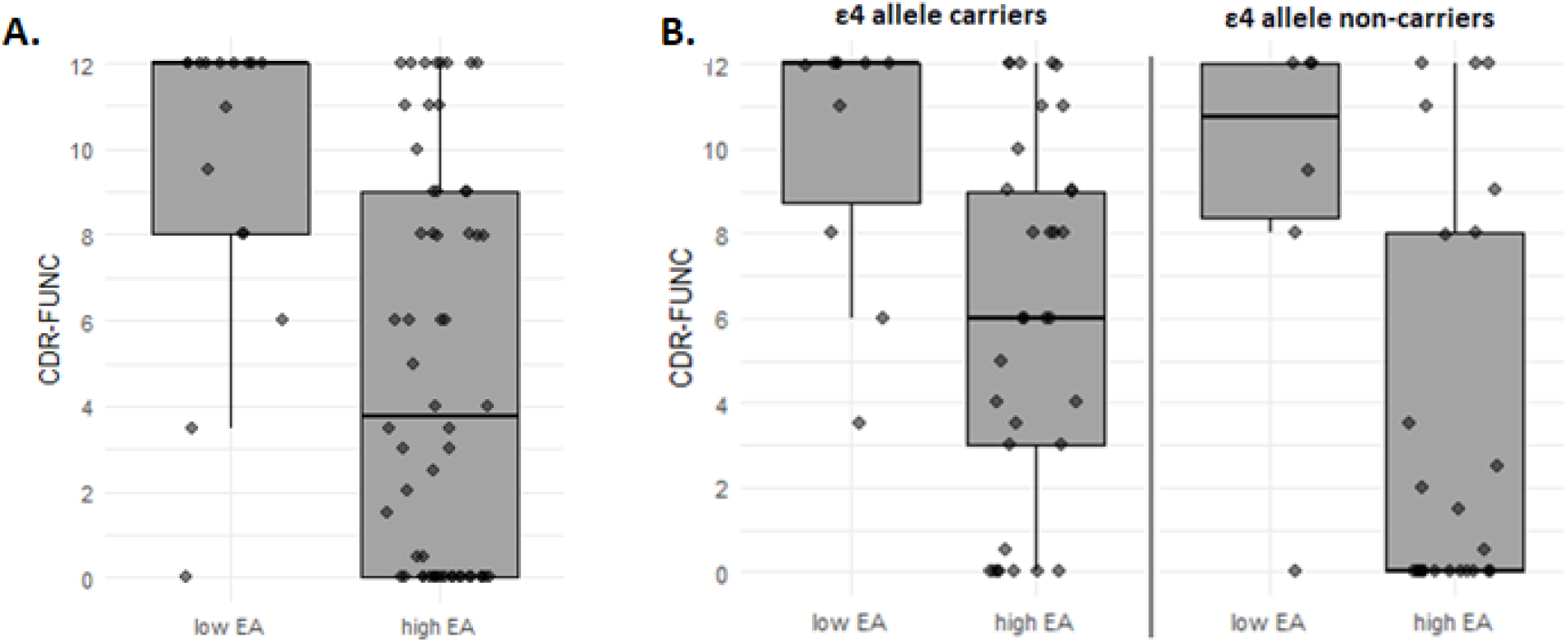
Box plot for the comparison between educational attainment (EA) levels (low and high) and CDR-FUNC score among advanced Alzheimer disease pathology individuals. (A) compare all individuals with the advanced Alzheimer disease pathology and (B) compare within subgroups stratified by the APOE ε4 allele.

To determine the effect of the *APOE* ε4 allele on the association between EA and CDR-FUNC in advanced log_10_(pTau181) individuals, we examined this relationship separately in ε4 carriers and non-carriers. We found that in both subgroups, individuals with low EA had significantly higher CDR-FUNC scores (ε4 allele carriers: median=12.0; Q1:8.75 - Q3:12.0; ε4 allele non-carriers: median=10.75, Q1:8.375-Q3:12) than those with high EA (ε4 allele carriers: median=6.0; Q1:3.0 - Q3:9.0; W=256.5, p=0.0008; ε4 allele non-carriers: median=0, Q1:0-Q3:8; W=117, p=0.029). However, the mean ranks of CDR-FUNC among high EA individuals were significantly higher in ε4 carriers compared to ε4 non-carriers (W=555.5, p=0.022). We did not observe a significant difference in CDR-FUNC score between ε4 carriers and non-carriers with low EA (W=33.5, p=0.719). Figure 1B shows the distribution of CDR-FUNC scores among advanced ADP individuals with low and high EA, stratified by ε4 allele.

## DISCUSSION

This study found that years of education are associated with better functional abilities in individuals with advanced log_10_(pTau181) level. This effect was observed in both ***APOE*** ε4 carriers and non-carriers, but the protective effect of high EA was stronger in non-carriers. These results extend our understanding of the protective effect that cognitive reserve and EA may have on the functional performance of individuals with AD pathology (Avila et al., 2021; Lovden et al., 2020; Seblova et al., 2020; Stern et al., 2020; Arenaza-Urquijo and Vemuri, 2018; Stern et al., 1994). Based on these findings, it is likely that increased years of education may explain why individuals with known genetic and AD biomarkers do not exhibit significant functional decline or impairment. It can be inferred that higher education can help individuals maintain their functional abilities even with AD pathology.

Our results highlight a complex relationship among the genetic risk factors, ADP-associated biomarkers, and cognitive reserve and their effect on cognitive and functional outcomes of AD. By investigating potential interactions among EA, ADP, and other genetic and modifiable risk and protective factors for AD, such as lifestyle factors (e.g., diet, exercise, sleep), or environmental exposures (e.g., air pollution, stress) we can enhance our understanding of the relative significance of these factors in the development of AD. This knowledge can then inform the design of comprehensive interventions aimed at preventing or delaying the onset of AD.

## Limitations

Findings from this study should be interpreted with caution considering several methodological and instrumentation limitations. First, the study population is relatively small and is nor representative of the global Black population. Therefore, we are unable to make universal claims that our findings will be observed in all individuals with African ancestry. Second, our operationalization of years in school as a proxy for educational attainment, does not capture quality of education, type of school, nature of school curriculum and location of school, all factors that could have impacted the findings. Third, the cross-sectional design of the data precluded us from making any causal claims in the associations between EA and functional status of individual with ADP. In spite of these limitations, the strong community-based representation of our sample, the heterogeneity of participants across multiple sites (Florida and North Carolina) addresses concerns about no having a representative sample, and the use of gold-standard instruments to assess target variables, as well as the public health and clinical value of our findings overshadow these limitations.

## Conclusion

In summary, this study shows the importance of using multiple variables such as genetics, education (an established social determinant of health), and AD biomarkers, to have a complete picture of the disease and its cognitive and functional outcomes. Notably, it demonstrates the need for comprehensive approaches to analyze the data, identify patterns associated with a risk of developing AD, and identify the specific combinations of factors most predictive of an individual’s risk of developing AD symptoms. Healthcare providers could use this information to identify individuals at high risk of AD and to develop personalized prevention and treatment plans for those individuals.

## Data Availability

Data are available through the National Institute on Aging Genetics of Alzheimers Disease Data Storage Site (NIAGADS) Data Sharing Service (DSS)

https://dss.niagads.org/datasets/ng00067/

